# Daily personalized music listening versus audiobook listening for chronic non-cancer pain in patients on long-term opioid therapy: a randomized controlled trial

**DOI:** 10.64898/2026.10.13.26365452

**Authors:** Jèssica Garrido-Pedrosa, María Fernanda Porto, Ancor Serrano-Afonso, Rocío Valenzuela, Lorena Zapata, María Florencia Vidal, Paloma Luna-Díaz, Emma Segura, Xim Cerda-Company, Sebastià Videla, Antoni Rodríguez-Fornells, Jennifer Grau-Sánchez, Víctor Fernández-Dueñas

## Abstract

Non-pharmacological adjuncts are needed for chronic non-cancer pain in patients receiving long-term opioid therapy. Music listening may reduce pain, but most previous trials have used passive controls, limiting attribution of effects specifically to music. In this open-label, parallel-group, proof-of-concept randomized controlled trial, adults with chronic non-cancer pain receiving long-term opioid therapy were randomized 1:1 to four weeks of daily home-based music or audiobook listening. Analgesic outcomes included pain intensity, responder rates, quantity of pain relief, and multidimensional pain assessment; secondary outcomes included mood, quality of life, global impression of change, and safety. Of 70 participants entering the run-in period, 56 were randomized (28 per group). Adjusting for baseline pain intensity in a linear mixed-effects model, the between-group difference at end of treatment was −1.26 NRS points (95% CI −1.98 to −0.55; p<0.001; d=−0.70), favoring music. A ≥30% pain-reduction criterion was met by 21.4% of participants in the music group and none in the audiobook group (p=0.023). An exploratory between-group difference in pain intensity favoring music was also observed at follow-up. Other analgesic and secondary outcomes showed no significant between-group differences, although exploratory within-group improvements in mood and health-related quality of life were observed with music. Both groups reported overall improvement in global perceived change, with ratings numerically favoring music. No intervention-related harms were observed. These findings support an analgesic benefit of music beyond the structured listening experience shared with the active comparator. Daily self-administered music listening may represent a feasible, low-cost adjunct for chronic non-cancer pain in patients receiving long-term opioid therapy.

## 1. Introduction

Chronic pain, defined as pain that persists or recurs for more than three months [55], affects more than a quarter of adults and is a leading cause of disability and reduced quality of life [11]. It frequently co-occurs with depression, anxiety, and sleep disturbance, and has been linked to substance misuse and premature mortality [8,24].

Many patients respond only partially to available therapies, and long-term opioid therapy remains commonly prescribed despite limited benefit and well-recognized risks, including tolerance, dependence, opioid use disorder, and opioid-induced hyperalgesia [12,23,26]. As chronic pain is increasingly understood as a multidimensional disorder shaped by biological, psychological, and social factors [36,47], management has shifted towards multimodal approaches, with growing interest in safe, scalable adjuncts that relieve symptoms without adding pharmacological burden [1,10].

Music listening has emerged as a promising therapy. Music, particularly when experienced as pleasurable, engages reward and pain-modulation systems and may also influence pain through attentional modulation, emotional regulation, and reward-related processes [31,34,44,53]. Music is inexpensive, accessible, easy to self-administer, and generally well accepted by patients [3,56]. Systematic reviews suggest that music-based interventions reduce pain and improve psychological outcomes in chronic pain [7,21,22,32].

However, evidence from well-controlled randomized trials remains limited. Most trials have compared music with passive controls (usual care, silence, or quiet rest), and much of the remaining work has compared different ways of delivering music rather than isolating the music effect itself [21,22]. Such designs cannot separate the effect of music from the non-specific effects of a structured listening activity, such as sustained attention and daily routine [19]. Active comparators that account for these features have rarely been used, leaving the specific analgesic contribution of music uncertain [2].

Clarifying the music-specific contribution is particularly relevant in patients on long-term opioid therapy. Chronic opioid exposure can alter reward processing, affective regulation, and pain modulation [18], systems also implicated in music-induced analgesia. Whether music confers a specific benefit beyond another structured, engaging listening activity in this context therefore remains unclear. Importantly, such a music-specific benefit would not necessarily imply a pain-specific mechanism. Music may preferentially engage reward and dopaminergic signaling, emotional regulation, positive affect, and attentional processes, which can in turn modulate the cognitive, affective, and motivational dimensions of pain experience [19,41]. A recent scoping review of mechanistic research identified convergent evidence for positive emotional valence, cognitive agency, and sensorimotor synchronization as candidate mechanisms, while also highlighting that most mechanistic studies have been conducted in experimental pain models in healthy volunteers [5]. In addition, few trials have examined outcomes beyond pain intensity, such as mood, quality of life, and opioid-related risk. Sex-related differences in treatment response have also rarely been addressed, despite well-documented sex differences in pain [48].

We therefore conducted a randomized controlled trial (RCT) comparing daily music listening with audiobook listening in adults with chronic non-cancer pain receiving long-term opioid therapy. Audiobook listening was chosen as an active comparator because it shares non-specific features with music (auditory stimulation, sustained engagement, and home-based daily practice) while lacking the musical component. We hypothesized that music listening reduces pain intensity more than audiobook listening.

## 2. Methods

### 2.1. Patient and public involvement

Patients were not involved in the design, conduct, or analysis of the trial. Following completion of the study, three individuals living with chronic pain contributed to the interpretation of the findings by providing feedback on their clinical relevance and applicability, as well as on their broader experience of living with chronic pain. Their contributions were considered when contextualizing the findings and developing dissemination materials.

### 2.2. Trial design

This was a single-center, two-arm, open-label, parallel-group, proof-of-concept RCT with a 1:1 allocation ratio. Individuals living with chronic non-cancer pain and receiving long-term opioid therapy were recruited at the Pain Unit of a tertiary hospital and randomly assigned to four weeks of structured daily music listening or audiobook listening, administered as an adjunct to their ongoing usual care and opioid therapy.

The study was approved by the Clinical Research Ethics Committee of Bellvitge University Hospital (UB-FCL-MUSIC 2022) and was conducted in accordance with the Declaration of Helsinki and applicable national regulations. Personal data were processed in accordance with EU Regulation 2016/679 (General Data Protection Regulation) and Spanish Organic Law 3/2018 on the protection of personal data and guarantee of digital rights. The trial was prospectively registered in ClinicalTrials.gov (identifier: NCT05726266), and the study design and procedures were described in the published protocol [25].

### 2.3. Protocol addenda

Two addenda to the published evaluation procedures were introduced to improve feasibility and linguistic and cultural appropriateness, without changing the outcome domains. First, pain intensity was recorded on the Numerical Rating Scale (NRS), a measure equivalent to the Visual Analogue Scale specified in the protocol but better suited to home self-administration and routine clinical use at the recruiting site. Second, multidimensional pain was assessed using the validated Spanish Pain Questionnaire (SPQ; *Cuestionario Español del Dolor*), the Spanish adaptation of the Short-Form McGill Pain Questionnaire, rather than the McGill Pain Questionnaire initially planned in the protocol, to ensure linguistic appropriateness for the study population.

### 2.4. Trial setting

The trial was conducted at the Pain Unit of Bellvitge University Hospital, a tertiary care hospital in Barcelona, Spain. All participants were managed as outpatients. Clinical assessments were conducted at the hospital, whereas the listening intervention was completed at home. During the intervention period, StopPain, a mobile application developed for the trial, was used for daily pain intensity recording and intervention monitoring.

### 2.5. Eligibility criteria

Eligible participants were adults (≥18 years) with a diagnosis of chronic non-cancer pain receiving a stable opioid treatment, with no changes in opioid prescription for at least four weeks before enrollment. All participants provided written informed consent before undergoing any study-specific procedures.

Exclusion criteria included pregnancy or lactation, and a history of organic brain disease, psychotic disorder, bipolar disorder, or intellectual disability. Participants were also excluded when the treating pain physician considered them at high risk of opioid use disorder or unlikely to comply with study procedures. Eligibility was assessed at Visit 1 and confirmed at Visit 2 before randomization.

### 2.6. Intervention and comparator

Participants in both groups were instructed to complete one self-administered listening session at home each day for 28 consecutive days. Sessions lasted between 30 and 60 minutes and were delivered through commercially available streaming platforms: Spotify for music listening and Audible for audiobook listening. Study-specific accounts with premium subscriptions were created for each participant to prevent commercial interruptions. Participants used their own smartphones and were instructed to listen using headphones in a quiet environment, alone, without carrying out other activities at the same time.

The interventions were delivered and monitored by two occupational therapists specialized in adult physical rehabilitation and with extensive experience in music-based interventions. The music-listening protocol and the structured musical-preference interview were developed by a qualified music therapist, who established the criteria for selecting and personalizing the listening material. Before enrollment, the two occupational therapists completed study-specific training on these procedures, the use of the StopPain application, and the strategies for promoting treatment fidelity and adherence.

Following randomization, each participant attended a structured interview with one of these occupational therapists to personalize the listening intervention. For participants allocated to music, therapists assessed musical preferences and created, in collaboration with the participant, individualized Spotify playlists that could be refined during the intervention. This personalized material was intended to be personally meaningful and enjoyable. For participants allocated to audiobooks, the therapist explored reading preferences and selected individualized Audible content to provide sustained auditory and cognitive engagement while avoiding the musical component hypothesized to contribute to analgesia.

The two interventions were matched for delivery format, session frequency and duration, home-based administration, personalization, therapist contact, and engagement in a structured listening activity. They differed primarily in the type of auditory content and whether music, the hypothesized active component, was present. Treatment fidelity was supported by weekly telephone contacts with the occupational therapist to sustain engagement, address technical difficulties, and adjust playlists or audiobook selections when necessary. No unplanned modifications to either intervention occurred during the trial. Participants continued their usual opioid regimen and other stable treatments throughout the study and were asked not to initiate any additional music-or audiobook-based intervention during the 28-day intervention period.

### 2.7. Study procedure

The study comprised three consecutive periods. During the 28-day pre-randomization run-in period, participants were enrolled but not yet randomized, and no listening intervention was administered; this period served to confirm stable opioid therapy and to characterize pain before randomization. The intervention period (28 days) began at randomization and comprised daily home-based listening. The follow-up period extended from the end-of-treatment (EoT) to the final end-of-study (EoS) assessment and included a safety telephone contact. Five assessments were scheduled across these periods, as detailed below; the schedule of outcome assessments is provided in **Figure S1**.

Visit 1: screening and start of the run-in period (Day −28 ± 3). Eligibility was initially assessed, written informed consent was obtained, and demographic and clinical data, including current opioid therapy, were collected. Participants received instructions on completing the daily paper diary. Throughout the run-in period, participants recorded daily pain-related information. An occupational therapist conducted weekly telephone calls to reinforce study procedures and encourage daily completion.

Visit 2: baseline assessment, eligibility confirmation, and randomization (Day 0). Eligibility was confirmed through review of the run-in data. Baseline outcome measures were assessed before randomization. Participants were then informed of their allocation, completed the structured personalization interview, and received training in the use of the StopPain application.

Intervention period (Days 1–28). Participants completed one home-based listening session each day and used the StopPain application for daily pain recording and adherence monitoring. Occupational therapists conducted weekly telephone contacts to support adherence and fidelity.

Visit 3: end-of-treatment assessment (Day 28 ± 3). All outcomes were assessed, adherence data were reviewed, and a safety evaluation was performed. Opioid therapy was reviewed and recorded. The protocol-controlled listening intervention ended at this visit. During the subsequent follow-up period, participants were not required to continue listening, but further listening was not restricted or recorded. Participants continued recording pain-related information in a paper diary until the final follow-up assessment.

Visit 4: telephone follow-up (Day 56 ± 5). Potential adverse events and current opioid treatment were recorded for safety monitoring. No outcome measures were collected.

Visit 5: end-of-study assessment (Day 84 ± 7). Participants completed the final, exploratory outcome assessment and concluded participation.

### 2.8. Outcomes

The outcome domains were prespecified in the published study protocol [25]; modifications to individual assessment instruments are described above. They were selected to capture clinically relevant dimensions of chronic non-cancer pain in individuals receiving opioid therapy.

#### Primary outcome domain: analgesic effect

The primary outcome domain comprised pain intensity on the 11-point Numerical Rating Scale (NRS; 0–10) [28], summed pain intensity difference (SPID), responder rates, and the Spanish Pain Questionnaire (SPQ). Detailed information on the SPQ is provided in the Supplementary Methods, whereas the derivation and analysis of SPID and responder status are described in Section 2.12.

#### Secondary outcomes

Secondary outcomes comprised anxiety and depressive symptoms (Hospital Anxiety and Depression Scale, HADS); mood (Profile of Mood States, POMS); self-esteem (Rosenberg Self-Esteem Scale, RSES); Patient Global Impression of Change (PGIC); health-related quality of life (36-item Short Form Health Survey, SF-36; and EuroQol five-dimension five-level questionnaire, EQ-5D-5L); opioid-misuse risk (Opioid Risk Tool, ORT; and Revised Screener and Opioid Assessment for Patients with Pain, SOAPP-R); and safety (UKU Side Effect Rating Scale). The schedule of outcome assessments is shown in **Figure S1**. Detailed descriptions of all outcome measures, including scoring procedures and psychometric properties, are provided in the Supplementary Methods.

### 2.9. Adverse events and harms

Adverse events were defined as any unfavorable medical occurrence arising during the study period, irrespective of suspected relationship to the assigned intervention. Harms were defined more specifically as adverse events attributable to the music or audiobook interventions (including the listening sessions, use of headphones, or the study application). Both were monitored throughout the study using active surveillance and participant self-report. Weekly telephone contacts with the occupational therapist allowed participants to be systematically asked about discomfort, emotional distress, worsening pain, and any other unexpected symptoms; daily self-reported data collected through the StopPain application provided complementary information. All reported adverse events were documented and classified according to severity, and their potential relationship to the intervention was assessed by the research team. Participants were instructed to discontinue a session if they experienced significant discomfort or distress and to report the event to the research team.

### 2.10. Sample size

This proof-of-concept trial was designed to provide an initial estimate of the treatment effect rather than a definitive assessment of efficacy. Consequently, the sample size was determined primarily on the basis of recruitment feasibility. A target sample of 70 participants (35 per group) was considered achievable within the planned recruitment period while providing sufficient precision to inform the design of future confirmatory trials. At the design stage, a complementary power calculation indicated that, assuming a repeated-measures design with at least three summary measurements per participant, a within-participant correlation of 0.30, a standard deviation of 4 NRS points, and a clinically meaningful between-group difference of 2 NRS points, a sample of 70 participants would provide approximately 82% power at a two-sided significance level of 5%.

### 2.11. Randomization: sequence generation, allocation, and blinding

The randomization sequence was computer-generated using permuted blocks of variable size (4 or 6) to ensure balanced allocation between groups. After eligibility had been re-assessed at Visit 2, the Pain Unit physician entered the participant’s details into a REDCap-based electronic case report form (eCRF) [27], which automatically assigned participants according to the pre-generated allocation sequence.

Allocation concealment was ensured through the centralized eCRF system, and outcome assessors did not have access to the random allocation sequence. Because of the behavioral nature of the interventions, blinding of participants and occupational therapists was not feasible, and the treating physician was also aware of allocation; however, all visit-based primary and secondary outcome assessments were administered by researchers who remained blinded to treatment allocation throughout the study. Participants assessed at Visit 1 but not randomized at Visit 2 were considered pre-randomization exclusions.

### 2.12. Statistical methods

Continuous variables were summarized as means and standard deviations for pre-randomization characteristics and as medians and interquartile ranges (IQR) for outcome variables; categorical variables were summarized as frequencies and percentages. Pre-randomization characteristics are presented descriptively by group, in line with recommendations for randomized trials [4].

The primary analysis followed the intention-to-treat (ITT) principle, with all randomized participants analyzed according to their assigned group. To assess potential differences associated with pre-randomization exclusion and post-randomization attrition, pre-randomization characteristics were compared between the ITT population and participants excluded before randomization, and between end-of-treatment completers and participants who discontinued after initiating the intervention, using Welch’s t test for continuous variables and Fisher’s exact test for categorical variables.

Analgesic efficacy was assessed from daily diary pain intensity together with the responder, SPID, and SPQ measures described below. Diary pain-intensity values were derived at each assessment as the participant-level mean of the NRS ratings over the corresponding three-day window (the first three run-in days for Screening [V1], the last three run-in days for Baseline [V2], the last three intervention days for EoT [V3], and the last three days before the end-of-study visit for Follow-up [V5]); for the few participants without evaluable diary data in a window, the NRS recorded at the corresponding clinical visit was used. A three-day average was used to provide a more stable estimate of pain intensity than a single-day measurement.

Pain intensity was compared between groups at EoT (V3) using the Mann–Whitney U test and analyzed longitudinally using two complementary linear mixed-effects models, each with a participant-specific random intercept. The between-group treatment effect was estimated with an analysis of covariance (ANCOVA) model in which the outcomes were pain intensity at EoT (V3) and Follow-up (V5), with fixed effects for treatment group, visit, and their interaction, and adjustment for Baseline (V2) pain intensity as a covariate. The longitudinal course of pain intensity (the within-group changes from Baseline (V2) and the overall Group × Visit interaction) was characterized with a second model that included Baseline (V2), EoT (V3), and Follow-up (V5) as repeated measures. Models were fitted using restricted maximum likelihood (REML), with Kenward-Roger degrees of freedom. Missing outcome data were accommodated within the mixed-effects framework under a missing-at-random assumption, so that all available observations contributed without imputation. The main comparison for pain intensity was the between-group difference at EoT (V3), calculated as the estimated marginal mean for the music group minus that for the audiobook group; negative values indicate lower pain intensity in the music group. The comparison at Follow-up (V5) was considered exploratory, because the protocol-controlled intervention ended at EoT (V3). Estimated marginal means were used to obtain between-group differences at each visit and within-group changes from Baseline (V2). Between-group contrasts are reported with nominal p values, whereas within-group contrasts were corrected for multiplicity using the Benjamini–Hochberg false discovery rate (FDR) procedure and are reported with both nominal p values and FDR-adjusted q values.

The robustness of the pain-intensity effect was assessed in several sensitivity analyses based on the same linear mixed-effects model. It was re-estimated with additional adjustment for sex, age, screening health-related quality of life (EQ-5D), and perceived social support (MSPSS). Effect modification by sex was examined by adding a sex × treatment interaction as an exploratory analysis. In addition, because music was administered as an adjunct to ongoing opioid therapy, it was examined whether the treatment effect was sensitive to the pre-randomization pharmacological regimen, adjusting the model sequentially for opioid step (major versus minor analgesic), number of concurrent drugs, and polypharmacy class.

The remaining outcomes in the primary analgesic domain were analyzed as follows. Quantity of pain relief was quantified using the summed pain intensity difference (SPID), calculated by subtracting each daily intervention-period NRS score from the participant’s baseline (V2) NRS score and summing these differences across the intervention period; higher values indicate greater pain relief. Under the ITT principle, participants without evaluable SPID data were assigned a value of 0 using baseline observation carried forward (BOCF), and groups were compared using the Mann–Whitney U test. A complete-case analysis restricted to participants with evaluable intervention-period diary data was performed as a sensitivity analysis, using Student’s t test where distributional assumptions were met. Responder status was operationalized from the daily NRS pain-intensity recordings collected during the intervention period. Participants were classified as responders if they showed a ≥30% reduction (and, separately, a ≥50% reduction) relative to baseline for at least three consecutive days; both thresholds are widely accepted as clinically meaningful [16]. Participants with insufficient evaluable data were conservatively classified as non-responders [54], and responder rates were compared between groups using Fisher’s exact test. The published protocol listed additional responder definitions based on maximum pain intensity over the previous 24 hours and rescue-medication use. These were not analyzed because the daily diary recorded a single NRS rating rather than a separate 24-hour maximum, and rescue-medication data were not captured in a format suitable for responder analysis. The Spanish Pain Questionnaire (SPQ) was compared between groups at EoT (V3) using the Mann–Whitney U test, with missing data imputed by BOCF under the ITT principle, and analyzed longitudinally using a linear mixed-effects model including the overall Group × Visit interaction.

Secondary continuous outcomes (HADS, POMS, Rosenberg Self-Esteem Scale, EQ-5D index, EQ-VAS, and SF-36) were compared between groups at EoT (V3) using the Mann–Whitney U test and analyzed longitudinally using linear mixed-effects models with participant-specific random intercepts. Within-group changes from Baseline (V2) to EoT (V3) for these outcomes were considered exploratory and were corrected for multiplicity using the Benjamini–Hochberg procedure within each group, with nominal and FDR-adjusted p values (q) reported. Safety and opioid-related continuous outcomes (UKU, ORT, and SOAPP-R) were similarly compared between groups at EoT (V3) using the Mann–Whitney U test and analyzed longitudinally using analogous mixed-effects models. For UKU, the Baseline (V2) assessment was used as the pre-treatment reference. ORT and SOAPP-R were assessed at Screening (V1), EoT (V3), and Follow-up (V5), and Screening (V1) therefore served as the pre-treatment reference for these measures. The Patient Global Impression of Change (PGIC), an ordinal outcome assessed at EoT (V3), was compared between groups using the Mann–Whitney U test; within each group, PGIC ratings were compared with the upper bound of the no-change range (“almost the same”, score = 2) using a one-sample Wilcoxon signed-rank test.

Effect sizes were calculated to aid interpretation. For model-based between-group contrasts, Cohen’s d was calculated by dividing the estimated between-group difference by the pooled baseline standard deviation. For within-group NRS changes, Cohen’s d was calculated by dividing the estimated change from Baseline (V2) by the corresponding group-specific baseline standard deviation, with positive values indicating improvement. For the complete-case SPID comparison, Cohen’s d was calculated as the difference in group means divided by the pooled standard deviation. The rank-biserial correlation was used for the PGIC. All statistical tests were two-sided, with nominal significance defined as p < 0.05 and, where correction for multiple testing was applied, FDR-adjusted q < 0.05. Analyses were performed using R version 4.5.2 (R Foundation for Statistical Computing) with the lme4, lmerTest, and emmeans packages.

## 3. Results

### 3.1. Participant flow diagram and pre-randomization characteristics

Participant flow is shown in **Figure 1**. Participant recruitment began on 18 May 2023, and the final follow-up assessment was completed on 16 October 2025. Seventy participants were enrolled at Visit 1 and entered the 28-day pre-randomization run-in period. Fourteen did not proceed to Visit 2 and were excluded before randomization and initiation of the intervention, most commonly because of insufficient run-in diary completion or disengagement during the run-in period. The protocol planned 35 randomized participants per group; however, these 14 pre-randomization exclusions resulted in a final randomized sample of 56 participants (28 per group).

**Figure 1.**
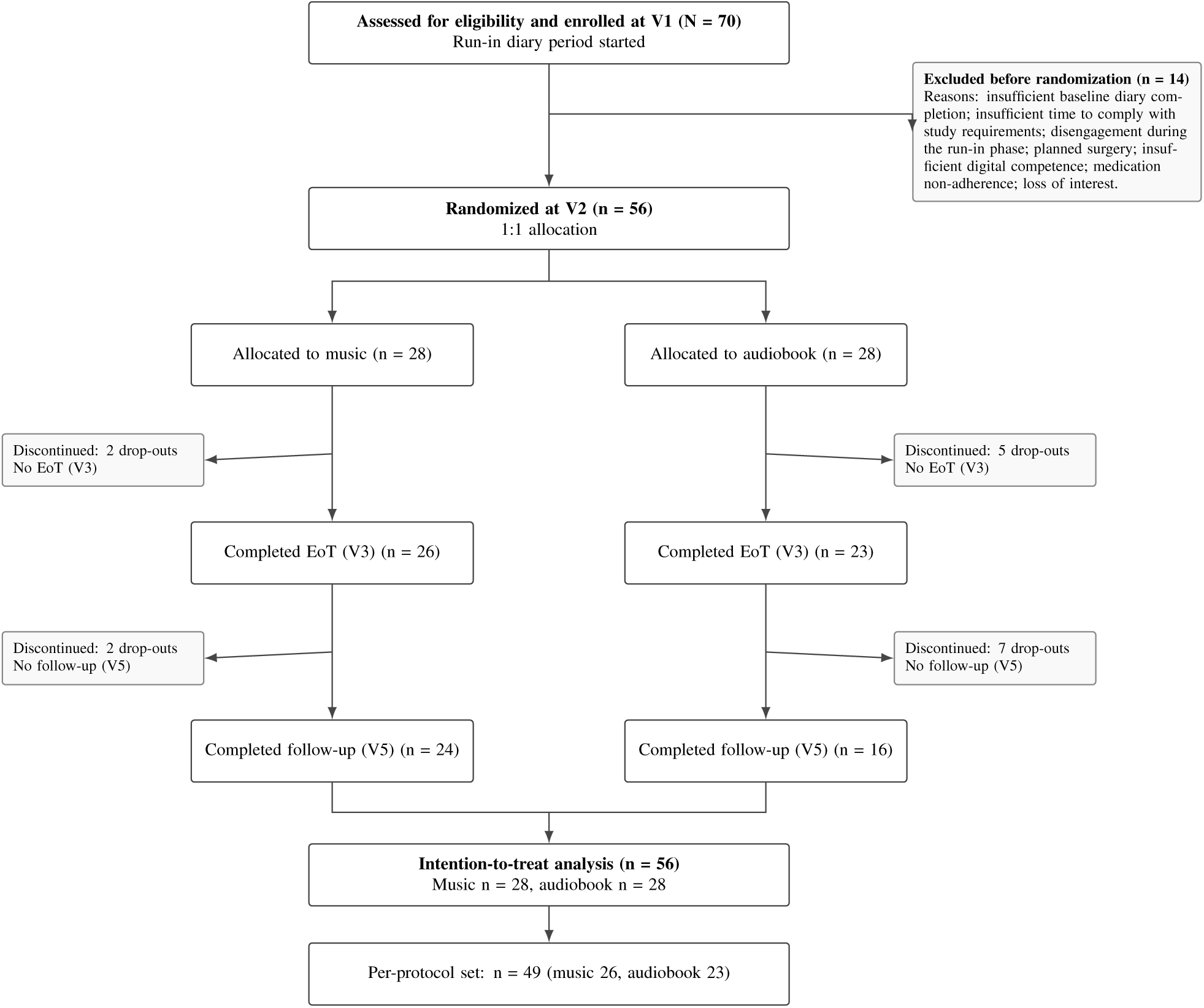
Participant flow (CONSORT diagram). Randomization occurred at the Baseline visit (V2); participants excluded before randomization are not part of the ITT population. *ITT*, intention-to-treat; *PP*, per-protocol; *V1*, screening; *V2*, Baseline; *V3*, end of treatment (EoT); *V5*, follow-up.

The ITT population comprised all 56 participants (28 music, 28 audiobook). All initiated the intervention. Forty-nine completed the end-of-treatment assessment (V3) (26 music, 23 audiobook), and 40 completed follow-up (V5).

Pre-randomization demographic and clinical characteristics were generally similar between groups (**Table 1**). The mean age of participants was 58.0 years, and 59% were women. Groups were comparable in educational level, employment status, pain characteristics and intensity, and sensitivity to musical reward, although perceived social support (MSPSS) was somewhat higher in the music group. Regarding opioid therapy, tramadol was the most common agent in both groups, while major opioids (i.e. fentanyl, tapentadol, morphine) were somewhat more frequent in the audiobook group. Opioid treatment remained stable throughout the intervention period in nearly all participants of both groups, with few and balanced changes (**Table S1**). Daily diary pain intensity during the run-in period was stable and comparable between groups, with no systematic trend before randomization (**Figure S2**).

**Table 1.** Pre-randomization characteristics of the intention-to-treat sample (n = 56).

|  | **Music (n = 28)** | **Audiobook (n = 28)** |
| --- | --- | --- |
| Age (years), mean (SD) | 58.3 (8.5) | 57.7 (11.9) |
| **Sex, n (%)** |  |  |
| Male | 10 (36) | 13 (46) |
| Female | 18 (64) | 15 (54) |
| **Education, n (%)** |  |  |
| Primary | 6 (21) | 6 (21) |
| Secondary | 9 (32) | 8 (29) |
| Vocational training | 9 (32) | 12 (43) |
| University | 3 (11) | 2 (7) |
| Master's | 1 (4) | 0 (0) |
| **Employment, n (%)** |  |  |
| Unemployed | 0 (0) | 1 (4) |
| Employed | 6 (21) | 3 (11) |
| Retired | 3 (11) | 10 (36) |
| Work disability | 19 (68) | 14 (50) |
| **Pain type, n (%)** |  |  |
| Chronic primary | 1 (4) | 1 (4) |
| Neuropathic | 10 (36) | 9 (32) |
| Musculoskeletal | 14 (50) | 17 (61) |
| Postsurgical/posttraumatic | 3 (11) | 1 (4) |
| **Pain severity at screening, n (%)** |  |  |
| Mild–moderate (0–6) | 10 (36) | 13 (46) |
| Moderate–severe (7–10) | 18 (64) | 15 (54) |
| **Opioid therapy, n (%)\*** |  |  |
| Fentanyl | 3 (11) | 6 (21) |
| Buprenorphine | 3 (11) | 2 (7) |
| Tramadol | 19 (68) | 16 (57) |
| Tapentadol | 2 (7) | 6 (21) |
| Oxycodone | 2 (7) | 2 (7) |
| Morphine | 0 (0) | 3 (11) |
| Social support (MSPSS), mean (SD) | 87.8 (10.0) | 77.7 (20.8) |
| Music reward (BMRQ), mean (SD) | 74.0 (12.6) | 72.5 (15.5) |

Values are mean (SD) for continuous variables and n (%) for categorical variables. \*Patients may receive more than one opioid; categories are not mutually exclusive. *BMRQ*, Barcelona Music Reward Questionnaire; *MSPSS*, Multidimensional Scale of Perceived Social Support; *SD*, standard deviation.

To assess potential differences associated with pre-randomization non-progression and post-randomization attrition, participants in the ITT population were compared with those who did not proceed to randomization and with those who discontinued after initiating the intervention. No statistically significant differences in measured pre-randomization characteristics were observed between participants who proceeded to randomization and those who did not (**Table S2**). Among participants who initiated treatment, those who completed the end-of-treatment assessment reported numerically higher pre-randomization perceived social support than those who discontinued, although this difference was not statistically significant (MSPSS: 85.0 vs 66.9; p = 0.130; **Table S3**).

### 3.2. Analgesic outcomes

#### Pain intensity (NRS)

At end of treatment (V3), observed pain intensity was numerically lower in the music group than in the audiobook group (median 6.1, IQR 4.1–7.0 vs 7.0, IQR 5.0–8.0; unadjusted between-group p = 0.092; **Table 2**). In the longitudinal linear mixed-effects model, adjusted for baseline pain intensity, the estimated between-group difference at EoT (V3) was −1.26 NRS points (95% CI −1.98 to −0.55; p = 0.0008; d = −0.70; **Figure 2A**), favoring music. From Baseline (V2) to EoT (V3), pain intensity decreased by 1.07 ± 0.26 NRS points in the music group (95% CI 0.44 to 1.70; q = 0.0002; d = 0.60), whereas no significant change was observed in the audiobook group (+0.24 ± 0.27 NRS points; q = 0.752; d = −0.14). The overall longitudinal pattern differed between groups (Group × Visit interaction χ²(2) = 12.98, p = 0.0015). At the exploratory follow-up assessment (V5), the model-estimated between-group difference continued to favor music (−0.90 NRS points, 95% CI −1.69 to −0.12; p = 0.024; d = −0.51; **Figure S3** and **Table S4**).

**Figure 2.**
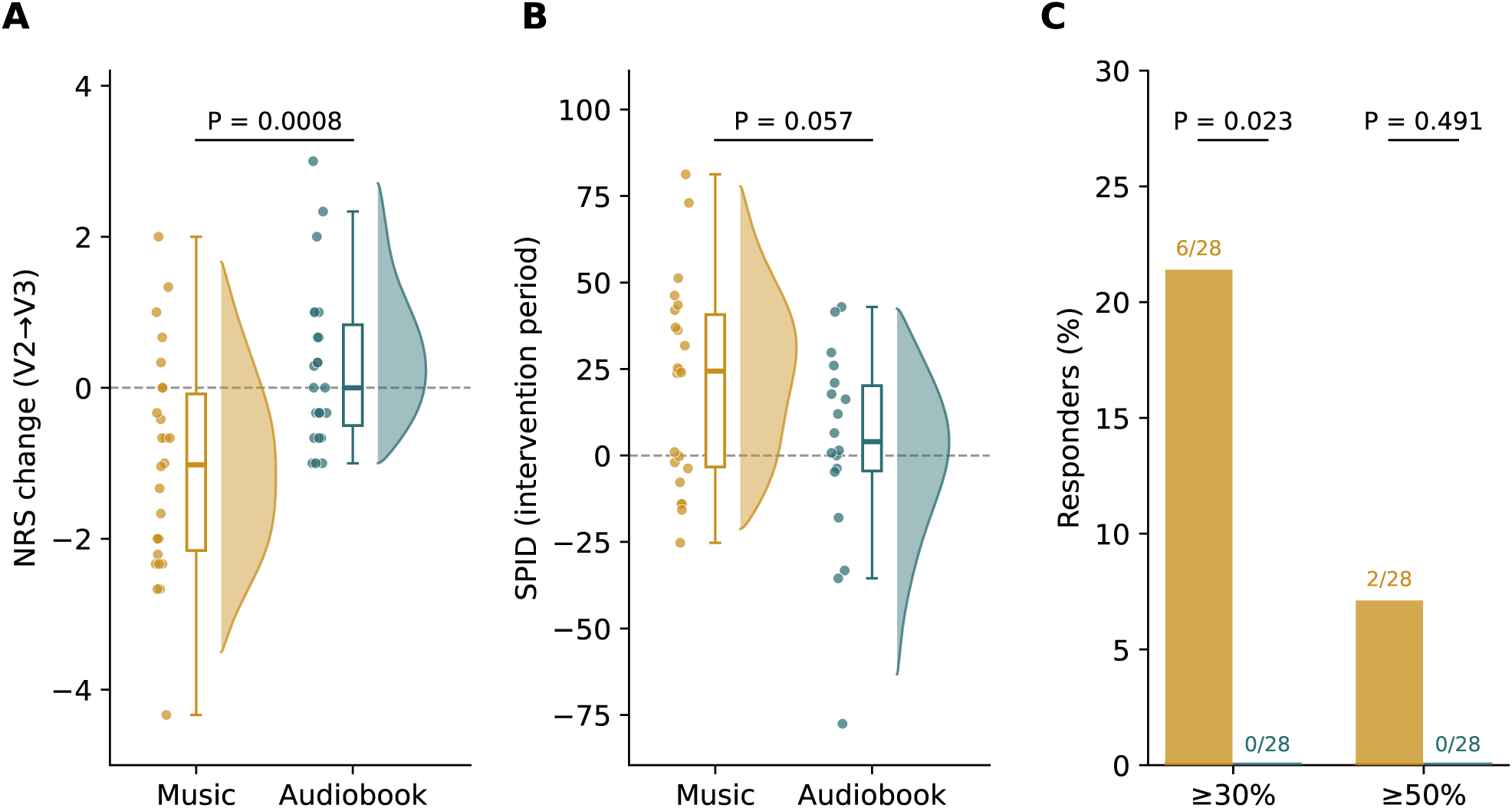
Analgesic outcomes. (A) Change in Numerical Rating Scale (NRS) pain intensity from Baseline (V2) to end of treatment (EoT, V3) by group; negative values indicate reduced pain. Individual participants are shown as points, boxes indicate the median and interquartile range, and half-violins show the distribution density. The p value corresponds to the baseline-adjusted between-group difference at EoT estimated using the linear mixed-effects model. (B) Summed pain intensity difference (SPID) over the intervention period; higher values indicate greater pain relief. The figure shows observed data from the 40 participants with evaluable intervention-period diaries, corresponding to the complete-case sensitivity analysis. Individual participants are shown as points, boxes indicate the median and interquartile range, and half-violins show the distribution density. Groups were compared using Student’s t-test. The ITT analysis with baseline observation carried forward for missing data is reported in Table 2. (C) Proportion of participants meeting the ≥30% and ≥50% responder criteria, with missing data conservatively classified as non-response. Labels indicate the number of responders among the 28 randomized participants per group. Groups were compared using Fisher’s exact test. Music is shown in mustard yellow and audiobook in petrol blue.

**Table 2.**
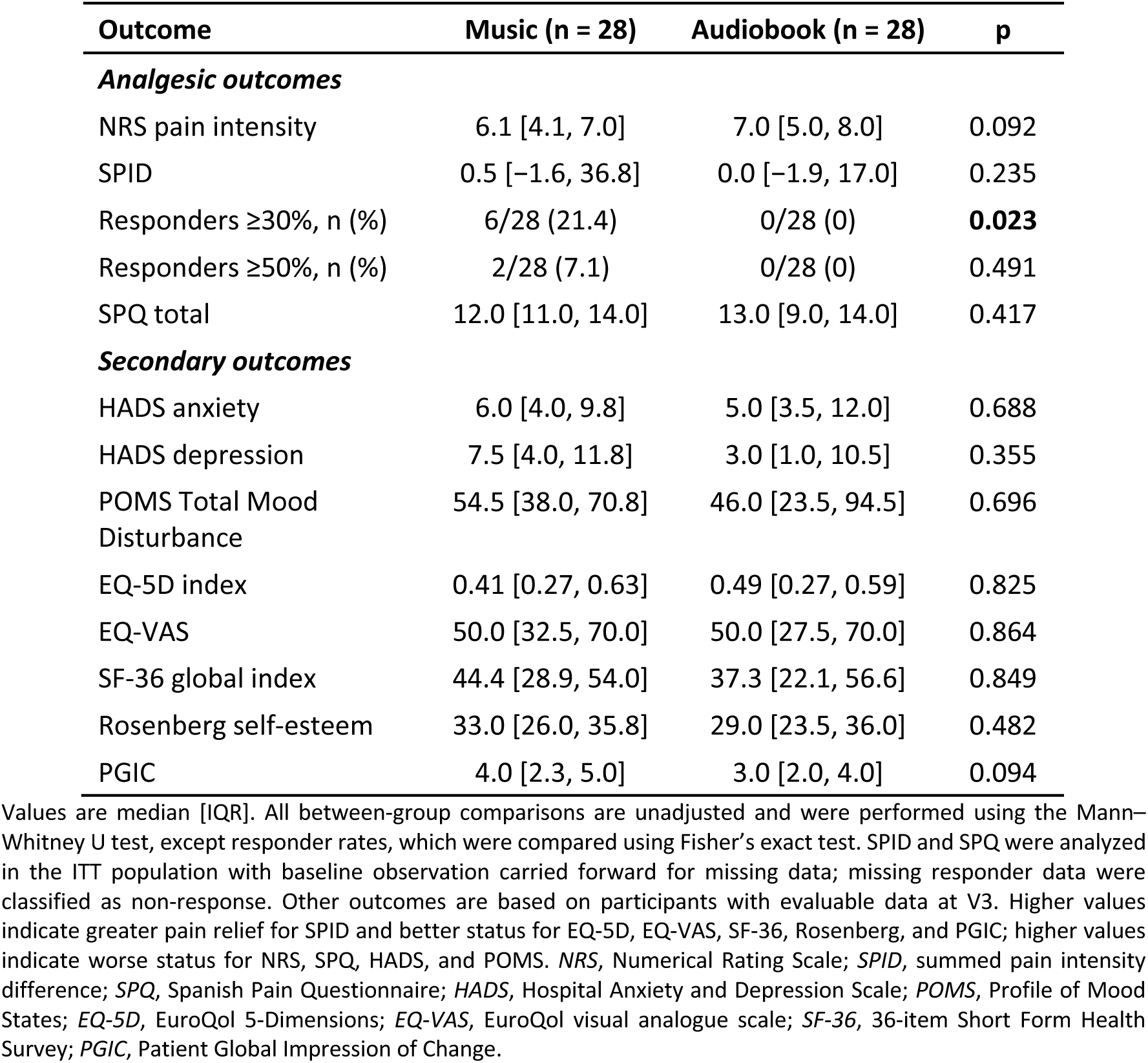
Outcomes at end of treatment by randomized group.

The model-based between-group difference at EoT was robust across sensitivity analyses. Adjustment for sex, age, health-related quality of life (EQ-5D), and perceived social support (MSPSS) yielded a slightly larger effect estimate (−1.43 NRS points, 95% CI −2.17 to −0.70; p = 0.0002; d = −0.80). There was no evidence that the treatment effect differed by sex (sex × treatment interaction, p = 0.536), although the trial was not powered to detect such interactions. Because music was administered alongside ongoing opioid therapy, we also examined whether the between-group estimate was sensitive to characteristics of the pre-randomization pharmacological regimen. Sequential adjustment for opioid step (major versus minor analgesic), number of concurrent drugs, and polypharmacy class left the estimate essentially unchanged (−1.25 to −1.26 NRS points, all p ≤ 0.001; **Table S5**).

#### Summed pain intensity difference (SPID)

In the ITT analysis using BOCF for missing data, SPID was higher in the music group (median 0.5, IQR −1.6 to 36.8) than in the audiobook group (median 0.0, IQR −1.9 to 17.0), although the between-group difference was not statistically significant (Mann–Whitney U, p = 0.235; **Table 2**). The low median values partly reflect the conservative BOCF approach, whereby participants without evaluable intervention-period diary data were assigned an SPID of 0, corresponding to no change from baseline. In the complete-case sensitivity analysis including the 40 participants with evaluable intervention-period diary data, mean SPID was 20.83 in the music group and 2.40 in the audiobook group (mean difference 18.43 points, 95% CI −0.63 to 37.50; Student’s t-test, p = 0.057; d = 0.62; **Figure 2B**).

#### Responder analyses

Using the analyzed responder criteria and non-responder imputation for missing data, 6 of 28 participants (21.4%) in the music group met the ≥30% responder criterion, compared with none of the 28 participants in the audiobook group (Fisher’s exact p = 0.023; **Figure 2C**). At the ≥50% threshold, 2 participants in the music group (7.1%) and none in the audiobook group met the responder criterion (p = 0.491).

#### Multidimensional pain (SPQ)

In the ITT analysis with BOCF for missing data, SPQ scores at EoT (V3) were similar between groups (music median 12.0, IQR 11.0–14.0 vs audiobook median 13.0, IQR 9.0–14.0; Mann–Whitney U, p = 0.417; **Table 2**). The adjusted linear mixed-effects model showed no significant between-group difference at EoT (adjusted difference +0.96 points, 95% CI −0.52 to 2.43; p = 0.200), and the Group × Visit interaction was also not significant (χ²(2) = 3.33, p = 0.190; **Figure 3A**).

**Figure 3.**
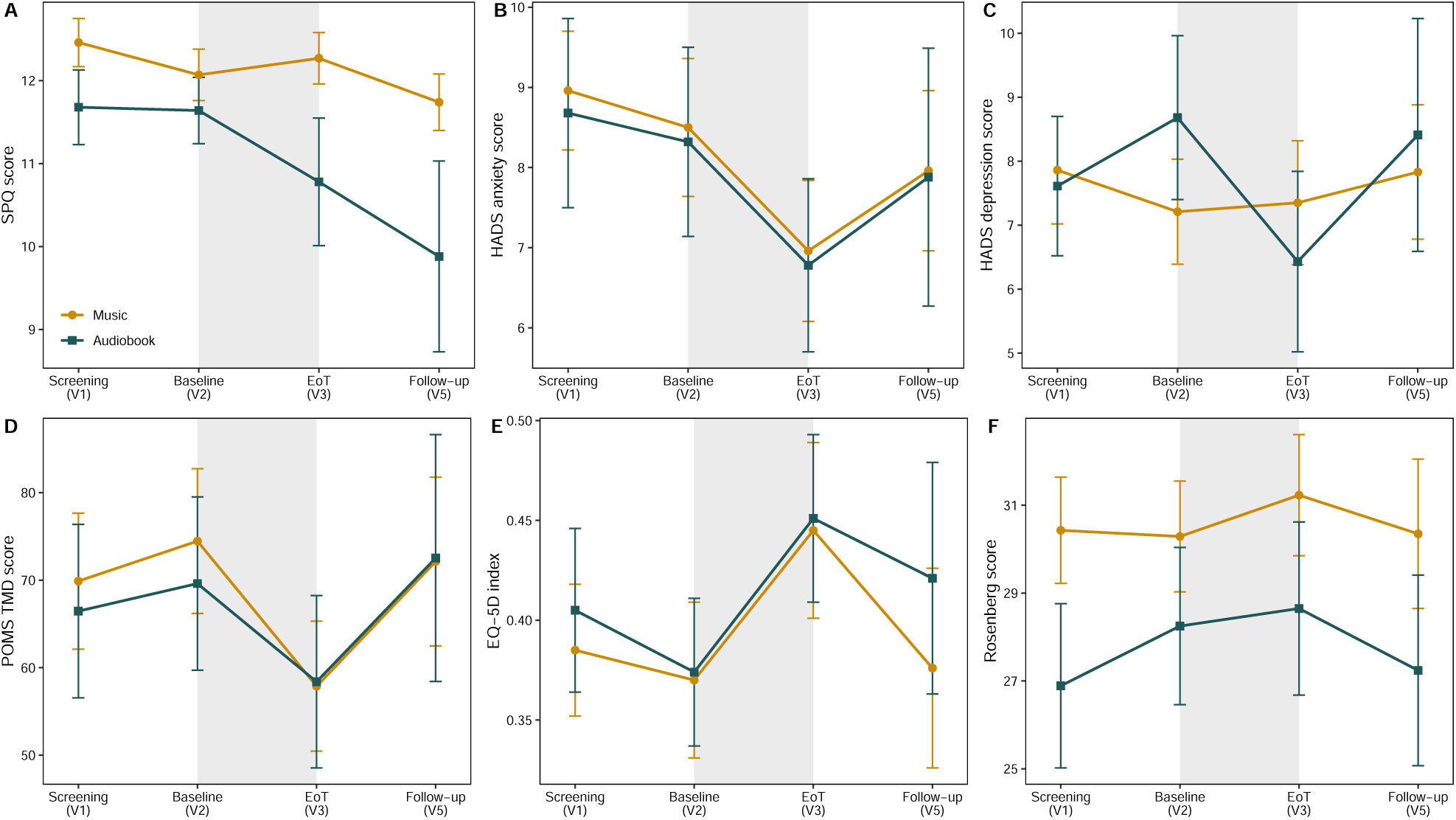
Multidimensional pain, mood, psychological, and health-related quality-of-life outcomes across study visits. (A) Spanish Pain Questionnaire (SPQ) total score. (B) Hospital Anxiety and Depression Scale (HADS) anxiety. (C) HADS depression. (D) Profile of Mood States (POMS) Total Mood Disturbance. (E) EQ-5D index. (F) Rosenberg Self-Esteem Scale. Data are shown across Screening (V1), Baseline (V2), end of treatment (EoT, V3), and Follow-up (V5). Points represent observed group means and ribbons indicate ±SEM; these trajectories are descriptive. The light gray shaded region marks the protocol-controlled intervention period from Baseline (V2) to EoT (V3); Follow-up (V5) was exploratory. Higher scores indicate worse status for SPQ, HADS, and POMS Total Mood Disturbance, and better status for EQ-5D and Rosenberg self-esteem. Music is shown in mustard yellow and audiobook in petrol blue.

### 3.3. Mood, psychological outcomes, and health-related quality of life

No secondary outcome showed a statistically significant between-group difference at EoT (V3) (**Table 2**). Exploratory within-group changes from Baseline (V2) to EoT (V3) are reported below; full results, including FDR-adjusted values, are provided in **Table S6**.

#### Hospital Anxiety and Depression Scale (HADS)

At EoT (V3), no significant between-group difference was observed for anxiety (music median 6.0, IQR 4.0–9.8 vs audiobook median 5.0, IQR 3.5–12.0; Mann–Whitney U, p = 0.688) or depression (music median 7.5, IQR 4.0–11.8 vs audiobook median 3.0, IQR 1.0–10.5; Mann–Whitney U, p = 0.355; **Table 2**). Descriptively, anxiety severity shifted toward lower HADS categories (abnormal → borderline → normal) more frequently in the music group: 9/26 participants improved by at least one category compared with 2/23 in the audiobook group. The number of participants in the abnormal anxiety category (score ≥11) decreased from 9 to 3 in the music group and from 10 to 7 in the audiobook group. Changes in depression categories showed no clear between-group pattern (music: 4 improved, 4 worsened; audiobook: 3 improved, none worsened). In the adjusted linear mixed-effects model, the between-group difference at EoT (V3) was not significant for anxiety (−0.50 points, 95% CI −2.41 to 1.41; p = 0.606) or depression (+1.85 points, 95% CI −0.29 to 3.99; p = 0.089), and the Group × Visit interaction was not significant for either subscale (anxiety χ²(2) = 0.29, p = 0.865, **Figure 3B**; depression χ²(2) = 3.71, p = 0.157, **Figure 3C**).

Exploratory within-group analyses from Baseline (V2) to EoT (V3) showed a reduction in anxiety in the music group that approached significance (−1.36 points, p = 0.055) but did not survive FDR correction (q = 0.096); no significant change was observed in the audiobook group. For depression, a reduction was observed in the audiobook group (−2.16 points, p = 0.009) that did not survive correction (q = 0.066), with no change in the music group (**Table S6**).

#### Profile of Mood States (POMS)

At EoT (V3), no significant between-group difference was observed in Total Mood Disturbance (music median 54.5, IQR 38.0–70.8 vs audiobook median 46.0, IQR 23.5–94.5; Mann–Whitney U, p = 0.696; **Table 2**), and no significant between-group differences were observed for any POMS subscale. In the adjusted linear mixed-effects model, the between-group difference at EoT (V3) was not significant (−8.06 points, 95% CI −22.08 to 5.96; p = 0.256), and the Group × Visit interaction was not significant (χ²(2) = 1.51, p = 0.471; **Figure 3D**). Exploratory within-group analysis showed a reduction in Total Mood Disturbance in the music group from Baseline (V2) to EoT (V3) (−16.87 points, p = 0.002) that remained significant after FDR correction (q = 0.013); no significant change was observed in the audiobook group (**Table S6**).

#### Health-related quality of life

At EoT (V3), no significant between-group difference was observed in the EQ-5D index (music median 0.41, IQR 0.27–0.63 vs audiobook median 0.49, IQR 0.27–0.59; Mann–Whitney U, p = 0.825; **Table 2**) or the EQ-VAS (music median 50.0, IQR 32.5– 70.0 vs audiobook median 50.0, IQR 27.5–70.0; Mann–Whitney U, p = 0.864; **Table 2**). In the adjusted linear mixed-effects model, the between-group differences at EoT (V3) were not significant (EQ-5D index +0.015, 95% CI −0.08 to 0.11, p = 0.764; EQ-VAS +3.44, 95% CI −7.55 to 14.43, p = 0.534), and the Group × Visit interaction was not significant (χ²(2) = 0.34, p = 0.843, **Figure 3E**; EQ-VAS χ²(2) = 0.67, p = 0.714). Exploratory within-group analysis showed an improvement in the EQ-5D index in the music group from Baseline (V2) to EoT (V3) (+0.075, p = 0.038), although it did not survive FDR correction (q = 0.088); no significant change was observed in the audiobook group (**Table S6**). EQ-VAS scores increased numerically in both groups (music: +6.16 points, p = 0.085; audiobook: +3.61 points, p = 0.407), but neither change reached statistical significance (**Table S6**).

For the SF-36 global index, no significant between-group difference was observed at EoT (V3) (music median 44.4, IQR 28.9–54.0 vs audiobook median 37.3, IQR 22.1–56.6; Mann–Whitney U, p = 0.849; **Table 2**; adjusted difference +3.02 points, 95% CI −3.44 to 9.47, p = 0.355), and the Group × Visit interaction was not significant (χ²(2) = 2.34, p = 0.311; **Figure S4A**). No significant between-group differences were observed across the eight SF-36 domains. Exploratory within-group analysis showed an improvement in the SF-36 global index in the music group from Baseline (V2) to EoT (V3) (+6.04 points, p = 0.008) that remained significant after FDR correction (q = 0.029); no significant change was observed in the audiobook group (**Table S6**).

#### Rosenberg Self-Esteem Scale

At EoT (V3), no significant between-group difference was observed in self-esteem (music median 33.0, IQR 26.0–35.8 vs audiobook median 29.0, IQR 23.5–36.0; Mann–Whitney U, p = 0.482; **Table 2**). In the adjusted linear mixed-effects model, the between-group difference at EoT (V3) was not significant (+1.50 points, 95% CI −1.09 to 4.08; p = 0.253), and the Group × Visit interaction was not significant (χ²(2) = 1.63, p = 0.443; **Figure 3F**). Exploratory within-group analyses showed no significant change from Baseline (V2) to EoT (V3) in either group (**Table S6**).

#### Patient Global Impression of Change (PGIC)

At EoT (V3), ratings were numerically higher in the music group (median 4.0, IQR 2.3–5.0) than in the audiobook group (median 3.0, IQR 2.0–4.0), although the between-group difference was not statistically significant (Mann–Whitney U = 381.5, p = 0.094; **Figure 4**). Within each group, PGIC ratings indicated improvement relative to the no-change category (both p ≤ 0.002).

**Figure 4.**
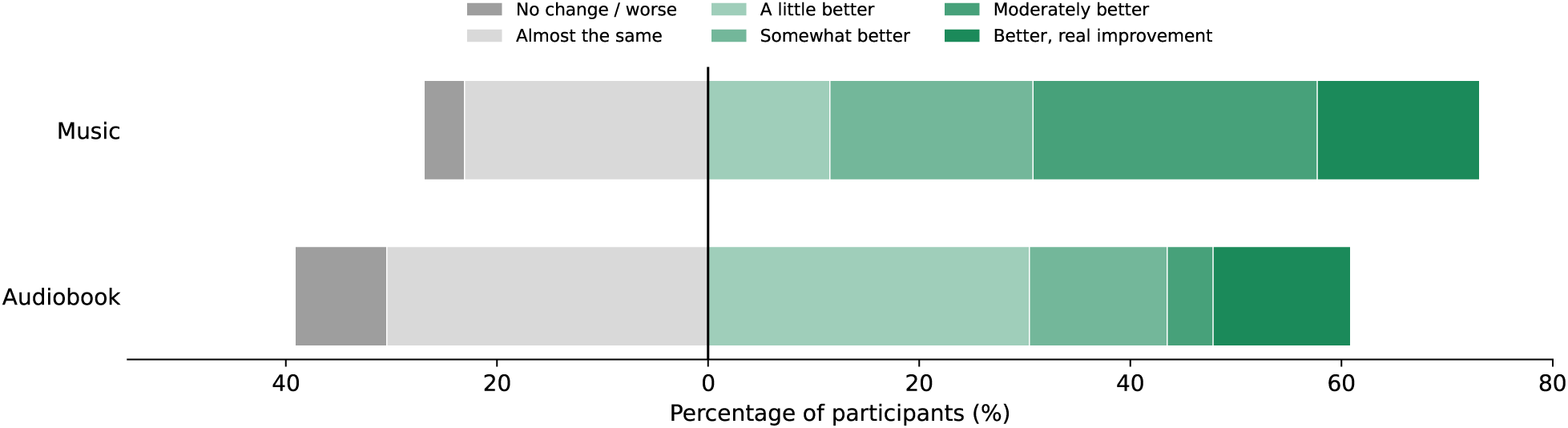
Patient Global Impression of Change (PGIC). Diverging stacked bars show the distribution of PGIC responses at end of treatment (EoT, V3) by group among participants who completed that visit. Bars are centered at the boundary between no change and improvement: grey segments to the left represent no change or worsening (“no change/worse” and “almost the same”), and green segments to the right represent increasing degrees of improvement. No participant selected the highest category (“much better”). Groups were compared using the Mann–Whitney U test, and responses within each group were compared with the upper bound of the no-change range (“almost the same”, score = 2) using a one-sample Wilcoxon signed-rank test.

### 3.4. Adverse events and harms

Adverse events and safety outcomes were monitored throughout the study. At EoT (V3), UKU total side-effect scores were numerically higher in the music group (median 10.0, IQR 5.5–14.8) than in the audiobook group (median 5.0, IQR 1.5–14.0), although a similar difference was already present at screening (V1 medians 11.5 vs 4.5). In the adjusted linear mixed-effects model, no significant between-group difference was observed at EoT (adjusted difference +1.69, 95% CI −2.45 to 5.83; p = 0.419; **Figure S4B** and **Table S7**).

Opioid-related risk measures did not increase in either group. ORT scores remained stable and comparable between groups (music median 1.5, IQR 1.0–5.0 vs audiobook 1.0, IQR 0.0–3.0 at EoT; adjusted difference +0.20, 95% CI −1.50 to 1.90; p = 0.818; **Table S7**). SOAPP-R scores decreased from screening to EoT in both groups (music median 16.0 to 8.0; audiobook 10.0 to 4.0). Scores were higher in the music group at screening (16.0 vs 10.0), but the adjusted between-group difference at EoT was not significant (+2.02, 95% CI −2.53 to 6.58; p = 0.386; **Table S7**).

No intervention-related harms were reported. Specifically, no participant reported discomfort, emotional distress, worsening pain, or other negative events related to music or audiobook listening, headphone use, or the study application, and no participant discontinued the intervention because of an intervention-related harm.

## 4. Discussion

This RCT evaluated whether daily music listening reduces pain intensity in patients with chronic non-cancer pain receiving long-term opioid therapy. To our knowledge, it is the first RCT to examine the analgesic benefit of music against an active, structured listening comparator in this population. At the end of the four-week intervention, the baseline-adjusted longitudinal analysis showed a moderate between-group difference in pain intensity favoring music, accompanied by a higher proportion of clinically meaningful pain responses. Other analgesic measures provided more limited support, with SPID favoring music but multidimensional pain scores showing little between-group separation. Because the audiobook comparator reproduced the main contextual and procedural features of the intervention, the overall pattern supports an analgesic benefit of music beyond participation in a structured listening intervention.

The magnitude and direction of the observed analgesic effect are broadly consistent with the existing literature. Meta-analyses have reported moderate reductions in pain with music-based interventions, with larger effects generally associated with repeated sessions, longer intervention periods, and participant-selected music [22]. Similar effects have been reported in systematic reviews focused specifically on chronic pain populations [7,21]. The effect observed here was of a similar magnitude to the pooled estimate from our recent systematic review (SMD ≈ −0.53), despite the use of an active comparator designed to control for non-specific effects of structured listening [21]. In our patient consultation, people living with chronic pain emphasized that even modest reductions in pain could be meaningful for daily functioning.

The secondary outcomes were broadly consistent with previous evidence. Between-group differences in anxiety, mood, and health-related quality of life were small and did not reach statistical significance. A recent meta-analysis reported a similar pattern, with more robust effects on pain and weaker, less consistent effects on anxiety and quality of life [7]. Notably, the only changes that remained significant after correction for multiplicity were observed in the music group, involving mood disturbance and general health status, whereas none survived correction in the audiobook group. Although exploratory and not evidence of a differential treatment effect, this pattern is compatible with broader effects of music on affective and health-related domains. This also aligns with feedback from our patient consultation, in which people with chronic pain described music and other immersive auditory activities as ways to divert attention from pain and remain engaged in daily life.

A key feature of this trial was the use of personalized audiobook listening as an active comparator. The audiobook reproduced important contextual and procedural features of the music intervention (personalization, sustained auditory engagement, home-based daily practice, and regular therapist contact) while lacking the musical features hypothesized to contribute to analgesia [17]. Achieving this balance remains a major methodological challenge in music-based intervention research [51]. Participants in both groups reported improvement on the PGIC, consistent with evidence that both music and spoken narratives can reduce pain relative to silence [42], and suggesting that structured listening may itself provide some benefit. Importantly, the between-group difference in pain intensity emerged against this active comparator, making shared components such as auditory stimulation, sustained engagement, and participation in a daily listening routine unlikely to fully account for the greater analgesic effect observed with music. Attentional mechanisms may nevertheless contribute, as music and spoken narratives engage attention differently [41].

The mechanisms underlying music-induced analgesia were not examined directly and can only be inferred from existing evidence. Among plausible music-related pathways, mesolimbic reward circuitry is strongly implicated in music-induced reward and pleasure [45,60], and pharmacological evidence supports a role for dopamine in musical reward [20]. Reward-related circuitry has also been implicated in music-induced analgesia [15,41]. Recent experimental work further suggests that the subjective experience of musical pleasantness may be particularly important: the effects of musical depth and arousal on pain were mediated by perceived pleasantness, supporting a role for affective valuation and reward rather than musical features alone [14]. Music-induced pleasure also engages endogenous opioid signaling [35,44,50], although its contribution to analgesia remains uncertain [40]. Importantly, music-induced hypoalgesia does not appear to be reducible to distraction. Recent experimental manipulation of attentional load showed additive effects of pleasant music and cognitive distraction on pain, with no evidence that the effect of music depended on attentional demand [13]. This is consistent with the benefit observed here against an active comparator requiring sustained auditory engagement. The exploratory improvements in mood and general health status observed with music are also compatible with affective and motivational contributions, although they do not establish mediation. Finally, the analgesic response observed in patients receiving stable long-term opioid therapy indicates that chronic opioid exposure did not preclude responsiveness to music in this sample. These pathways likely interact and cannot be disentangled in the present study.

This trial has several methodological strengths. It focused on a clinically relevant and understudied population of patients with chronic non-cancer pain receiving long-term opioid therapy, for whom safe non-pharmacological adjuncts are particularly needed. Analyses followed the intention-to-treat principle, with all randomized participants analyzed according to their assigned group and outcome-specific approaches to missing data. Pain intensity was assessed prospectively through daily diaries and at scheduled study visits, reducing reliance on retrospective recall. The intervention was standardized and delivered according to a prespecified protocol, with individualized content selected through structured interviews. The between-group estimate for pain intensity was robust to adjustment for pre-randomization covariates, including perceived social support, and for characteristics of the pharmacological regimen. Although major opioids were somewhat more frequent in the audiobook group before randomization, these adjustments left the estimated effect essentially unchanged, suggesting that the observed difference was not driven by baseline pharmacological imbalance. Neither intervention was associated with reported harm.

Some limitations should be acknowledged. This was a single-center proof-of-concept trial with a modest sample size and was not designed to detect small-to-moderate differences across secondary outcomes or exploratory subgroups. The sample also included participants with very long-standing pain, in some cases exceeding 10 years, but the available sample size precluded examining whether pain duration influenced treatment response. Although 70 participants entered the run-in period, 14 did not proceed to randomization, resulting in 56 randomized participants rather than the 70 planned in the protocol and reducing the precision of the estimates. This limited precision may partly explain why the unadjusted cross-sectional comparison of pain intensity at EoT, although directionally consistent, did not reach statistical significance, whereas the baseline-adjusted longitudinal model showed a moderate between-group difference favoring music. The open-label design was unavoidable, although visit-based outcome assessments were conducted by blinded assessors and an active comparator was used to minimize bias. However, the audiobook comparator could not ensure equivalence between conditions in reward value, pleasantness, or emotional engagement, factors that may themselves contribute to analgesia. Follow-up was limited to eight weeks after treatment, and post-treatment listening was neither restricted nor systematically recorded, so continued music listening may have contributed to the difference observed at follow-up. Finally, the mediation analysis involving opioid treatment described in the published protocol could not be performed because structured longitudinal opioid-dose data were unavailable. However, sensitivity analyses adjusting for characteristics of the pre-randomization pharmacological regimen left the between-group pain-intensity estimate essentially unchanged.

Important questions remain open. The treatment effect did not vary significantly by sex, although the trial was underpowered to detect interaction effects. This remains relevant because sex is an important source of variability in pain, and women constituted 59% of the sample. Individual differences in sensitivity to musical reward may also influence treatment response, suggesting that future trials should examine whether interventions can be better tailored to individual reward profiles [45]. Whether music is particularly beneficial for patients at higher risk of opioid misuse could not be reliably assessed here and remains an important question for larger trials. The relative merits of self-selected and researcher-selected music also remain debated, although emerging evidence suggests that subjective responses to the music, particularly pleasantness and emotional engagement, may be important determinants of hypoalgesia [14,22,31]. Future studies should also examine how the format of delivery shapes benefit. In our patient consultation, participants valued peer and group-based approaches, reflecting a need for recognition and social support that is well documented in chronic pain populations [9].

In summary, four weeks of personalized daily music listening produced a reduction in pain intensity in patients with chronic non-cancer pain receiving long-term opioid therapy. The effect was observed against an active audiobook comparator and was not accompanied by evidence of harm or increased opioid-related risk, supporting an analgesic benefit of music beyond the structured listening experience shared by both groups. As a low-cost, home-based, and self-administered intervention, daily music listening may represent a feasible adjunct to multidisciplinary pain management in a population with limited non-pharmacological options. Larger multicenter trials with longer follow-up are needed to establish the durability and generalizability of the effect, assess potential opioid-sparing benefits, and clarify the mechanisms involved.

## Data Availability

The data and analysis code generated during this study are available from the corresponding author upon reasonable request, under a data transfer agreement where required to protect participant confidentiality.

## Acknowledgements

We thank the members of the Patient Expert Committee, Juan Carlos Gonsalves, Angela Irigoyen, and Carla Pastor, for discussing the study results with us and providing valuable comments and perspectives that helped inform the interpretation of the findings. We also thank undergraduate student Laura Guzmán for her help with participant supervision. Finally, we especially thank Basetis S.A. for developing the StopPain app at a reduced cost.

This work was supported by Neuroscience Program, IDIBELL-Bellvitge Institute for Biomedical Research (21VAR007); Plan Nacional Sobre Drogas, Ministerio de Sanidad (2021I068, 2025I011); Ministerio de Ciencia e Innovación, Agencia Estatal de Investigación (CEX2021-001159-M).

During the preparation of this manuscript, the authors used Claude (Anthropic) and ChatGPT (OpenAI) to assist with language editing and statistical code review. All AI-assisted content was reviewed and edited by the authors, who take full responsibility for the final content of the manuscript.

The authors declare no competing interests.

## Author contributions

Conceptualization: S.V., A.R.-F., J.G.-S., V.F.-D.; Methodology: E.S., S.V., A.R.-F., J.G.-S., V.F.-D.; Investigation: J.G.-P., M.F.P., A.S.-A., R.V., L.Z., M.F.V., P.L.-D., J.G.-S.; Data curation: J.G.-P., M.F.P., M.F.V., R.V.; Formal analysis: E.S., X.C.-C., V.F.-D.; Writing – original draft: J.G.-P., J.G.-S., V.F.-D.; Writing – review & editing: J.G.-P., M.F.P., A.S.-A., R.V., L.Z., M.F.V., P.L.-D., E.S., X.C.-C., S.V., A.R.-F., J.G.-S., V.F.-D.; Supervision and project administration: J.G.-S., V.F.-D.; Funding acquisition: S.V., A.R.-F., V.F.-D. All authors approved the final version of the manuscript and agreed to be accountable for all aspects of the work.

## Notes

### Competing Interest Statement

The authors have declared no competing interest.

### Clinical Trial

NCT05726266

### Author Declarations

The Clinical Research Ethics Committee of Bellvitge University Hospital gave ethical approval for this work (reference: UB-FCL-MUSIC 2022).

