## Supplementary data for "Daily personalized music listening versus audiobook listening for chronic non-cancer pain in patients on long-term opioid therapy: a randomized controlled trial"

### Supplementary Methods

This section provides detailed descriptions of the outcome measures summarized in the main text (Section 2.8, Outcomes). The outcome domains were prespecified in the published study protocol [25]; differences in the individual assessment instruments are described in Section 2.3.

#### 1. Multidimensional pain assessment

##### 1.1. Spanish Pain Questionnaire (SPQ)

Multidimensional pain was assessed using the Spanish Pain Questionnaire (SPQ) [38], the validated Spanish adaptation of the McGill Pain Questionnaire (MPQ) [46], was administered. The total pain intensity score ranges from 0 to 14 and is derived from three dimensions: sensory (0–7), affective (0–6), and evaluative (0–1), with higher scores indicating greater pain intensity.

#### 2. Emotional status and mood

##### 2.1. Hospital Anxiety and Depression Scale (HADS)

Emotional status was assessed using the Hospital Anxiety and Depression Scale (HADS), a widely used instrument composed of two subscales, one for depression and the other for anxiety. Each subscale consists of seven items scored from 0 (best possible) to 3 (worst possible), and the subscale score is obtained by summing the item scores. Scores from 0 to 7 are considered “normal”, 8 to 10 “borderline”, and 11 to 21 “abnormal”. Participants can therefore be classified as improved, unchanged, or worsened according to movement to a lower, identical, or higher severity category. This instrument has demonstrated good internal consistency, with Cronbach’s alpha coefficients above 0.80 [37,61].

##### 2.2. Profile of Mood States (POMS)

Mood was evaluated using the Profile of Mood States (POMS) [49]. This self-report inventory consists of 65 items assessing six domains: fatigue–inertia, anger–hostility, vigor–activity, confusion–bewilderment, depression–dejection, and tension–anxiety. Participants rate each item on a 5-point Likert scale reflecting their current affective state, from 0 ("not at all") to 4 ("extremely"). The Total Mood Disturbance (TMD) score is obtained by summing the five negative subscales and subtracting the vigor–activity score; higher TMD scores therefore indicate worse mood and lower scores a more positive mood state.

#### 3. Self-esteem

##### 3.1. Rosenberg Self-Esteem Scale (RSES)

Self-esteem was assessed using the Rosenberg Self-Esteem Scale (RSES) [52], in its validated Spanish version [43]. This 10-item self-report scale measures global self-worth through positively and negatively worded statements, each rated on a 4-point Likert scale (1 to 4). Total scores range from 10 to 40, with higher scores indicating greater self-esteem.

#### 4. Global impression of change

##### 4.1. Patient Global Impression of Change (PGIC)

The Patient Global Impression of Change (PGIC) [33] is a patient-reported outcome measure that captures an individual’s perceived overall improvement over the course of treatment. This 7-point Likert scale provides a global evaluation encompassing multiple domains, including pain, physical functioning, emotional wellbeing, and daily activities. Scores range from 1 ("no change or worse") to 7 ("much better"), with higher scores reflecting greater perceived clinical improvement.

#### 5. Health-related quality of life

##### 5.1. 36-item Short Form Health Survey (SF-36)

Health-related quality of life was assessed using the 36-item Short Form Health Survey (SF-36) [58], in a version validated for Spanish-speaking populations [57]. The instrument evaluates eight health domains: Physical Functioning, Physical Role, Bodily Pain, General Health, Vitality, Social Functioning, Emotional Role, and Mental Health. Domain scores range from 0 (maximum disability) to 100 (no disability), with higher scores indicating better health status. The eight domains were analyzed individually and summarized as a global index, calculated as the unweighted mean of the eight domain scores.

#### 5.2. EuroQol 5-dimensions 5-level (EQ-5D-5L)

To complement this assessment, the EuroQol 5-dimensions 5-level (EQ-5D-5L) questionnaire was used [29,30]. This instrument evaluates five dimensions (mobility, self-care, usual activities, pain/discomfort, and anxiety/depression), each rated across five severity levels from 1 (no problems) to 5 (extreme problems). Responses were converted into a single health utility index using the Spanish value set [30], where higher scores represent better health-related quality of life. Participants also provided a self-rated health state on a visual analogue scale (EQ-VAS) ranging from 0 (worst imaginable health) to 100 (best imaginable health).

### 6. Safety and opioid-related outcomes

#### 6.1. Opioid Risk Tool (ORT)

The risk of opioid misuse was assessed using the Opioid Risk Tool (ORT), a self-administered screening instrument for patients prescribed opioids for chronic pain [59]. The ORT considers personal and family histories of substance misuse, age, history of preadolescent sexual abuse, and specific psychiatric comorbidities. Total scores range from 0 to 26, categorizing patients as low risk (0–3), moderate risk (4–7), or high risk (≥8); higher scores indicate a greater probability of future opioid misuse.

#### 6.2. Revised Screener and Opioid Assessment for Patients with Pain (SOAPP-R)

The Revised Screener and Opioid Assessment for Patients with Pain (SOAPP-R) [6] was used to evaluate the risk of opioid misuse in patients undergoing long-term opioid therapy. It comprises 24 self-report items identifying behaviors and characteristics associated with potential misuse, each rated on a 5-point Likert scale from 0 ("never") to 4 ("very often"). Total scores range from 0 to 96, with higher scores indicating greater risk; a cut-off score of ≥ 18 indicates an elevated risk of aberrant medication-related behavior.

#### 6.3. UKU Side Effect Rating Scale

Adverse effects were assessed using the UKU Side Effect Rating Scale [39]. This scale includes 48 items grouped into four dimensions (psychic, neurological, autonomic, and other side effects), each scored from 0 ("absent") to 3 ("severe"), allowing a detailed assessment of the type and intensity of side effects.

**Table S1.** Opioid treatment stability during the intervention and follow-up periods, by group.

| Period / treatment change | Music | Audiobook |
|---|---|---|
| **Intervention period (up to end of treatment)** | **(n = 26)** | **(n = 23)** |
| No change | 24 | 21 |
| Dose increase | 1 | 0 |
| Dose decrease | 0 | 0 |
| Drug switch | 0 | 1 |
| Unclear | 1 | 1 |
| **Follow-up period (up to end of study)** | **(n = 24)** | **(n = 16)** |
| No change | 17 | 11 |
| Dose increase | 1 | 0 |
| Dose decrease | 5 | 3 |
| Drug switch | 0 | 1 |
| Unclear | 1 | 1 |

Values are numbers of participants assessed at each visit. Changes in opioid treatment were classified from physician medication reports. Denominators correspond to participants who completed the end-of-treatment and end-of-study assessments, respectively.

**Table S2.** Attrition analysis A: comparison of the intention-to-treat population (n = 56) with participants excluded before the randomization (n = 14).

| | **ITT (n = 56)** | **Excluded (n = 14)** | ***p*** |
|---|---|---|---|
| Age (years), mean (SD) | 58.0 (10.3) | 59.4 (9.6) | 0.647 |
| **Sex, n (%)** | | | 0.225 |
| Male | 23 (41) | 3 (21) | |
| Female | 33 (59) | 11 (79) | |
| **Education, n (%)** | | | 0.088 |
| No formal education | 0 (0) | 1 (7) | |
| Primary | 12 (21) | 6 (43) | |
| Secondary | 17 (30) | 1 (7) | |
| Vocational training | 21 (38) | 6 (43) | |
| University | 5 (9) | 0 (0) | |
| Master’s | 1 (2) | 0 (0) | |
| Social support (MSPSS), mean (SD) | 82.7 (17.0) | 76.5 (22.7) | 0.363 |

Values are mean (SD) or n (%); *p* values from Welch’s *t*-test (continuous) and Fisher’s exact test (categorical). *ITT*, intention-to-treat; *MSPSS*, Multidimensional Scale of Perceived Social Support.

**Table S3.** Attrition analysis B: comparison of per-protocol completers (n = 49) with participants who discontinued after treatment initiation (n = 7).

| | **Completers (n = 49)** | **Dropouts (n = 7)** | ***p*** |
|---|---|---|---|
| Age (years), mean (SD) | 58.1 (9.7) | 57.0 (14.8) | 0.849 |
| **Sex, n (%)** | | | 0.429 |
| Male | 19 (39) | 4 (57) | |
| Female | 30 (61) | 3 (43) | |
| **Education, n (%)** | | | 0.380 |
| No formal education | 0 (0) | 0 (0) | |
| Primary | 9 (18) | 3 (43) | |
| Secondary | 14 (29) | 3 (43) | |
| Vocational training | 20 (41) | 1 (14) | |
| University | 5 (10) | 0 (0) | |
| Master's | 1 (2) | 0 (0) | |
| Social support (MSPSS), mean (SD) | 85.0 (14.0) | 66.9 (27.1) | 0.130 |

Values are mean (SD) or n (%); *p* values from Welch's *t*-test (continuous) and Fisher's exact test (categorical). *MSPSS*, Multidimensional Scale of Perceived Social Support.

**Table S4.** Numerical Rating Scale (NRS) pain intensity by group and visit, with model-estimated between-group differences at end of treatment and follow-up.

| Visit | Music, mean (SEM) | Audiobook, mean (SEM) | Between-group [95% CI], *p* |
|---|---|---|---|
| Screening (V1) | 7.06 (0.31) | 6.72 (0.35) | — |
| Baseline (V2) | 6.84 (0.36) | 6.56 (0.32) | — |
| Post (V3, EoT) | 5.71 (0.40) | 6.71 (0.34) | −1.26 [−1.98, −0.55], **0.0008** |
| Follow-up (V5) | 5.89 (0.42) | 6.65 (0.49) | −0.90 [−1.69, −0.12], **0.024** |

Means and SEM are observed values. Screening (V1) is shown descriptively and was not included as an outcome observation in the models. Between-group differences (music minus audiobook) at EoT (V3) and Follow-up (V5) are model-estimated adjusted contrasts from the linear mixed-effects analysis of covariance, which included treatment group, visit, their interaction, Baseline (V2) pain intensity as a covariate, and a participant-specific random intercept. No inferential between-group comparison was performed at Baseline (V2). Negative estimates indicate lower pain intensity in the music group. *CI*, confidence interval; *EoT*, end of treatment; *SEM*, standard error of the mean.

**Table S5.** Sensitivity of the NRS between-group effect at end of treatment (V3) to adjustment for the pre-randomization pharmacological regimen.

| **Model** | **Estimate [95% CI]** | ***p*** |
|---|---|---|
| M0: Main model | −1.26 [−1.98, −0.55] | **0.0008** |
| M1: + opioid step (major vs minor) | −1.25 [−1.98, −0.52] | **0.0011** |
| M2: + opioid step + number of drugs | −1.26 [−2.00, −0.53] | **0.0010** |
| M3: + opioid step + drugs + polypharmacy | −1.26 [−2.01, −0.52] | **0.0012** |

Estimates (music minus audiobook) at end of treatment (V3) are from the linear mixed-effects analysis of covariance (adjusted for Baseline [V2] pain intensity), sequentially adding opioid step (major vs minor analgesic), number of concurrent drugs, and polypharmacy class as covariates. *CI*, confidence interval; *NRS*, Numerical Rating Scale.

**Table S6.** Within-group changes (baseline to end of treatment) for secondary outcomes, by group.

| Outcome | Music: estimate (*p*) | q (FDR) | Audiobook: estimate (*p*) | q (FDR) |
|---|---|---|---|---|
| HADS anxiety | −1.36 (0.055) | 0.096 | −0.81 (0.274) | 0.384 |
| HADS depression | −0.03 (0.969) | 0.969 | −2.16 (0.009) | 0.066 |
| POMS Total Mood Disturbance | −16.87 (0.002) | **0.013** | −7.80 (0.165) | 0.288 |
| EQ-5D index | +0.075 (0.038) | 0.088 | +0.061 (0.104) | 0.288 |
| EQ-VAS | +6.16 (0.085) | 0.119 | +3.61 (0.407) | 0.475 |
| SF-36 global index | +6.04 (0.008) | **0.029** | +3.39 (0.156) | 0.288 |
| Rosenberg self-esteem | +1.38 (0.117) | 0.136 | +0.17 (0.853) | 0.853 |

Within-group changes from Baseline (V2) to end of treatment (V3), estimated from the trajectory linear mixed-effects models. Negative values indicate a reduction; higher scores indicate worse status for HADS and POMS, and better status for EQ-5D, EQ-VAS, SF-36, and Rosenberg. The q column gives the Benjamini–Hochberg FDR-adjusted value within each group. *FDR*, false discovery rate.

**Table S7.** Safety and opioid-related risk outcomes at screening, end of treatment, and follow-up, by group.

| Outcome / Visit | Music | Audiobook | *p* |
|---|---|---|---|
| ***UKU total score*** | | | |
| Screening (V1) | 11.5 [4.8, 16.5] | 4.5 [2.0, 13.2] | 0.106 |
| EoT (V3) | 10.0 [5.5, 14.8] | 5.0 [1.5, 14.0] | **0.039** |
| Follow-up (V5) | 6.0 [3.5, 14.5] | 4.0 [0.0, 9.0] | 0.165 |
| ***ORT*** | | | |
| Screening (V1) | 1.5 [1.0, 5.0] | 1.0 [0.0, 4.0] | 0.261 |
| EoT (V3) | 1.5 [1.0, 5.0] | 1.0 [0.0, 3.0] | 0.264 |
| Follow-up (V5) | 2.0 [0.5, 6.5] | 1.0 [0.0, 4.0] | 0.410 |
| ***SOAPP-R*** | | | |
| Screening (V1) | 16.0 [6.8, 20.2] | 10.0 [2.0, 19.8] | 0.184 |
| EoT (V3) | 8.0 [4.2, 12.8] | 4.0 [0.0, 9.5] | 0.073 |
| Follow-up (V5) | 5.0 [2.0, 13.0] | 2.0 [0.0, 5.0] | 0.202 |

Values are median [IQR] among participants with evaluable data at each visit. p values are from unadjusted between-group comparisons using the Mann–Whitney U test at each visit. Higher scores indicate greater side-effect burden (UKU) or greater risk of opioid misuse (ORT, SOAPP-R). ORT and SOAPP-R were assessed at Screening (V1), EoT (V3), and Follow-up (V5); for UKU, Screening (V1) values are shown for consistency across scales. Model-adjusted between-group differences at EoT are reported in Section 3.4. EoT, end of treatment; IQR, interquartile range; ORT, Opioid Risk Tool; SOAPP-R, Revised Screener and Opioid Assessment for Patients with Pain; UKU, UKU Side Effect Rating Scale.

**Figure S1.** Study design: visit schedule and outcome assessment timeline.

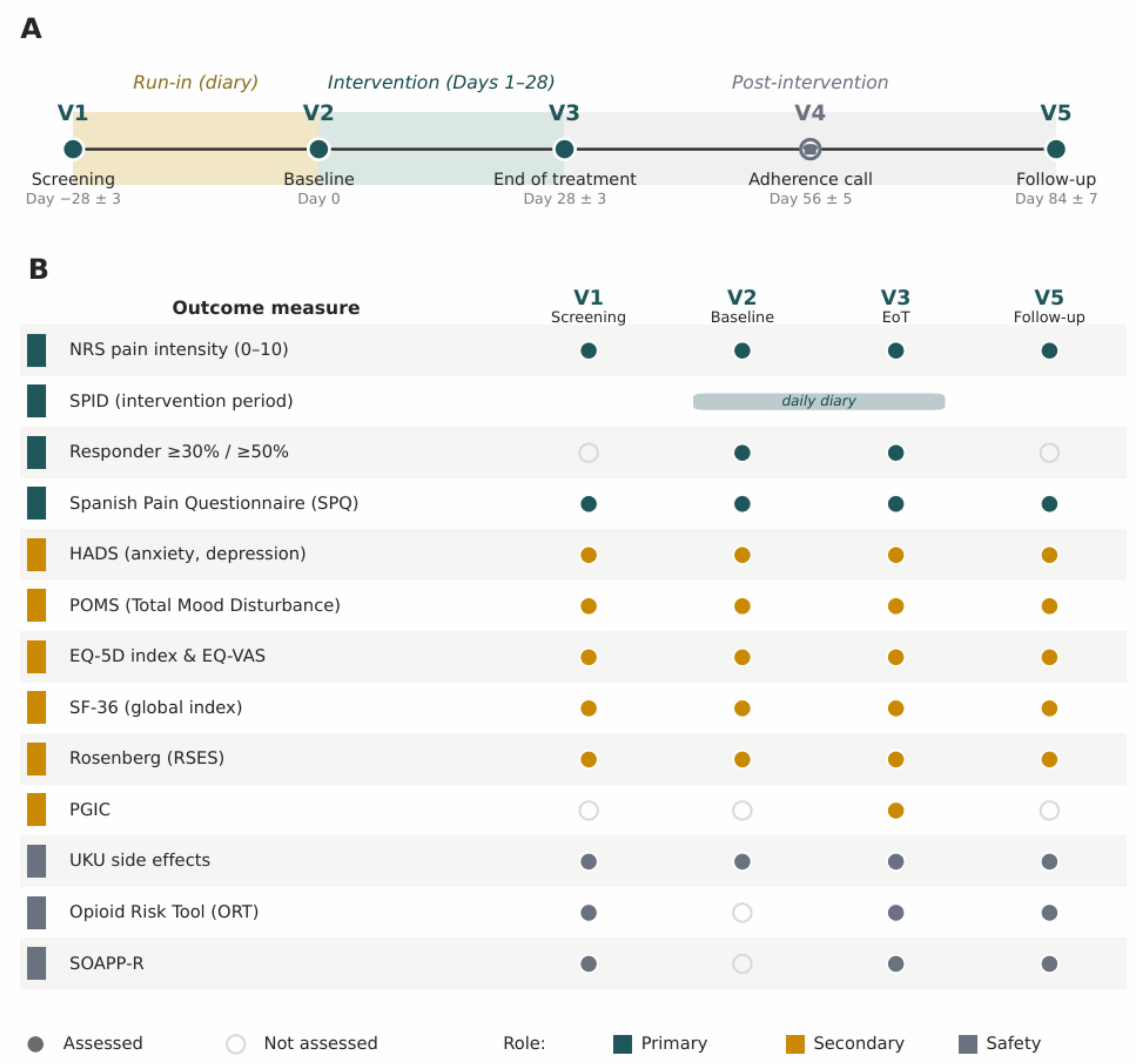


(A) Study visits and timing (days). Visit 1 (Screening, day −28 ± 3): start of the run-in diary period; Visit 2 (Baseline, day 0): start of the 28-day intervention; Visit 3 (End of treatment, day 28 ± 3); Visit 4 (day 56 ± 5): adherence and safety telephone call, with no outcome assessment; Visit 5 (Follow-up, day 84 ± 7): end-of-study assessment. Shaded bands indicate the run-in, intervention, and post-intervention phases. (B) Outcome measures and assessment schedule (filled circle, assessed; open circle, not assessed). Summed pain intensity difference (SPID) was derived from daily diary data collected during the intervention period (V2–V3). Marker color indicates outcome domain: analgesic (petrol blue), secondary (mustard yellow), or safety (grey). *NRS*, Numerical Rating Scale; *SPID*, summed pain intensity difference; *SPQ*, Spanish Pain Questionnaire; *HADS*, Hospital Anxiety and Depression Scale; *POMS*, Profile of Mood States; *EQ-5D*, EuroQol 5-Dimensions; *EQ-VAS*, EuroQol visual analogue scale; *SF-36*, 36-item Short Form Health Survey; *RSES*, Rosenberg Self-Esteem Scale; *PGIC*, Patient Global Impression of Change; *ORT*, Opioid Risk Tool; *SOAPP-R*, Revised Screener and Opioid Assessment for Patients with Pain.

**Figure S2.** Daily pain intensity during the run-in period, by group.

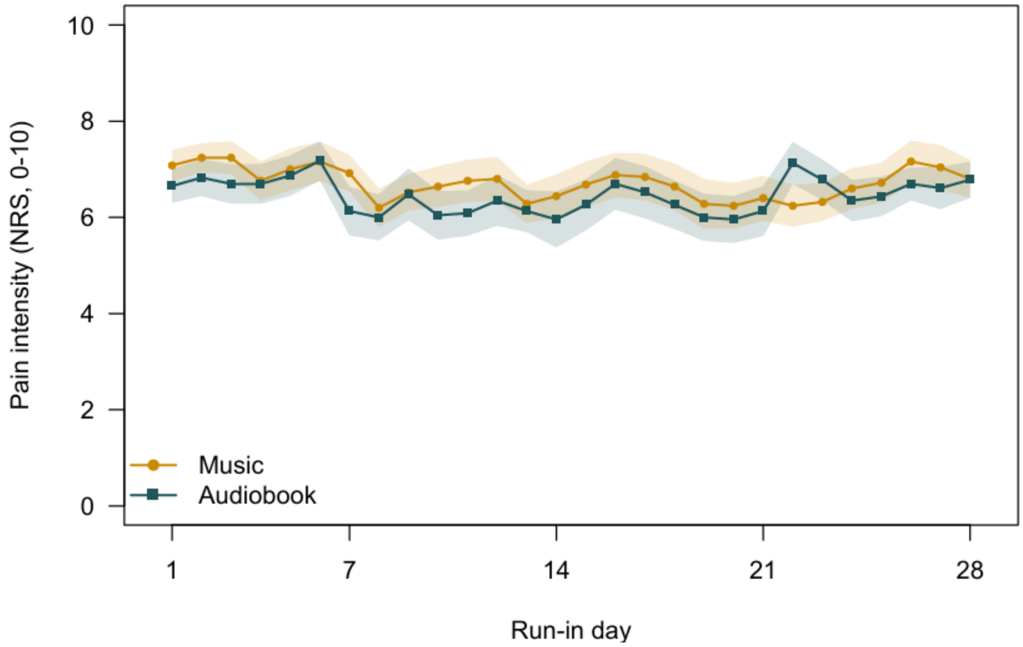


Daily pain intensity (NRS, 0–10) during the 28-day pre-randomization run-in period among participants with evaluable diary data, grouped according to subsequent randomized allocation. Points represent daily group means and shaded bands indicate ±SEM; lines connect consecutive daily means. Higher scores indicate greater pain intensity. Music is shown in mustard yellow and audiobook in petrol blue.

**Figure S3.** Numerical Rating Scale (NRS) pain intensity over the study and change at follow-up.

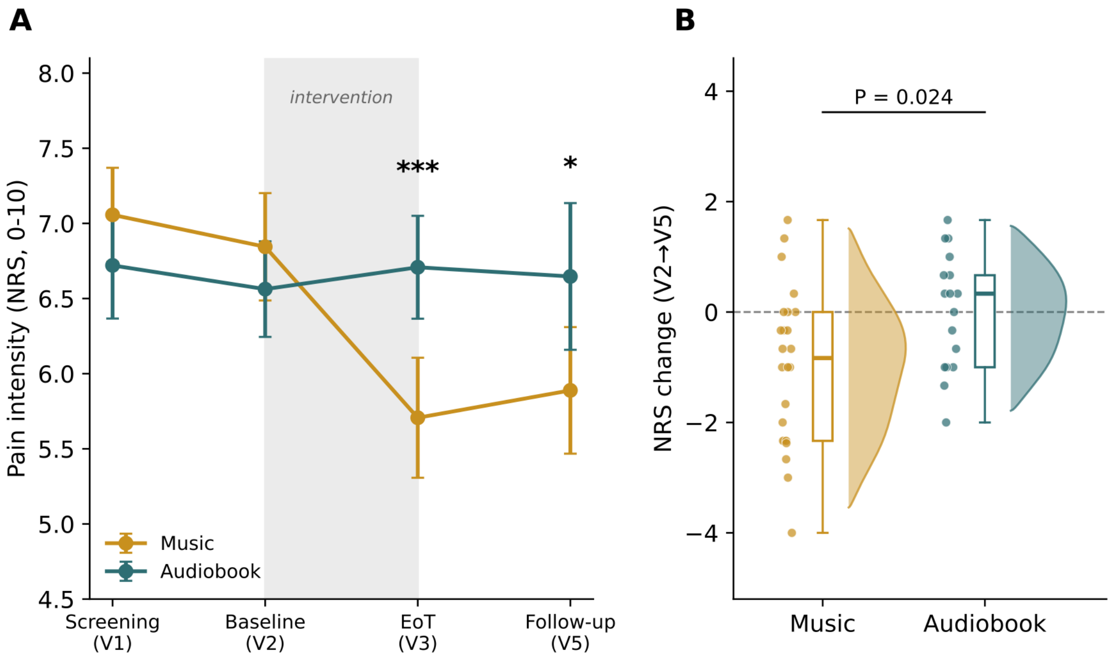


(A) NRS pain intensity (0–10) across Screening (V1), Baseline (V2), end of treatment (EoT, V3), and Follow-up (V5). Points represent observed group means and error bars indicate ±SEM. Visit-level NRS values were derived from participant-level diary averages: the first three run-in days for Screening (V1), the last three run-in days for Baseline (V2), the last three intervention days for EoT (V3), and the last three available diary days before Follow-up (V5); see Methods. The shaded region marks the protocol-controlled intervention period (V2–V3). Baseline-adjusted between-group differences at EoT (V3) and Follow-up (V5) were estimated using the linear mixed-effects model; ***$p < 0.001$, *$p < 0.05$. (B) Raincloud plot of individual change in NRS pain intensity from Baseline (V2) to Follow-up (V5); negative values indicate reduced pain. Points represent individual participants, boxes indicate the median and interquartile range, and the half-violin shows the distribution density. Exploratory between-group difference at Follow-up was estimated using the baseline-adjusted linear mixed-effects model. Music is shown in mustard yellow and audiobook in petrol blue.

**Figure S4.** Additional outcomes over the four study visits by group.

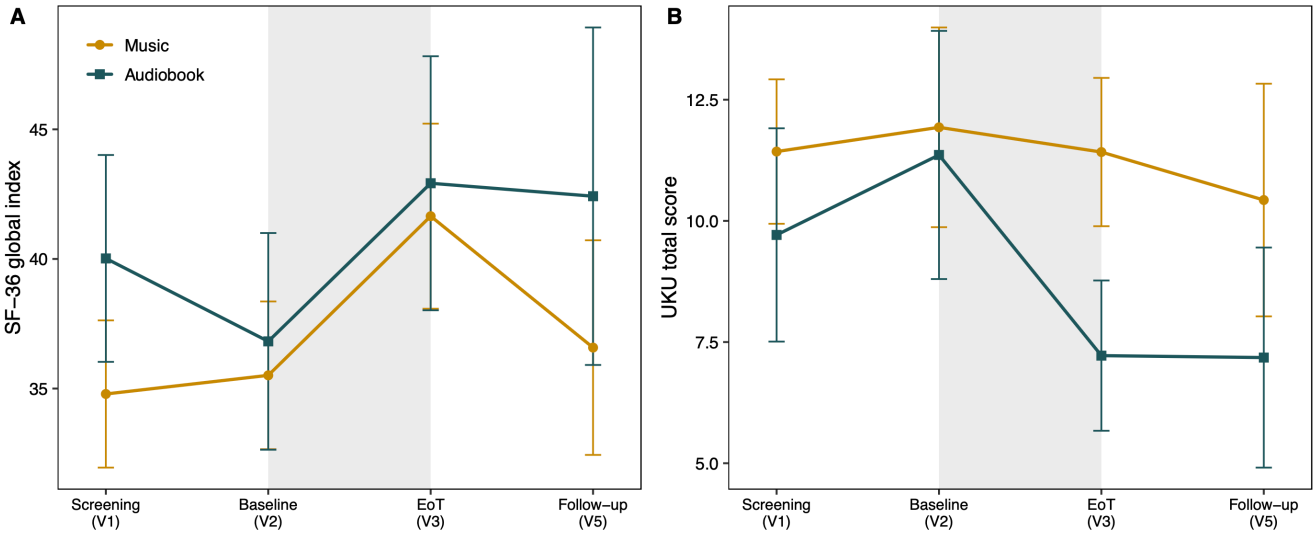


(A) SF-36 global index (unweighted mean of the eight domain scores, 0–100); higher scores indicate better health status. (B) UKU total side-effect score; higher scores indicate greater side-effect burden. Data are shown as observed group means ±SEM across Screening (V1), Baseline (V2), end of treatment (EoT, V3), and Follow-up (V5). The light gray shaded region marks the protocol-controlled intervention period from Baseline (V2) to EoT (V3). No between-group significance markers are shown. Music is shown in mustard yellow and audiobook in petrol blue.
